# Observer: Creation of a Novel Multimodal Dataset for Outpatient Care Research

**DOI:** 10.1101/2025.05.18.25327837

**Authors:** Kevin B. Johnson, Basam Alasaly, Kuk Jin Jang, Eric Eaton, Sriharsha Mopidevi, Ross Koppel

## Abstract

**Objective:** To support ambulatory care innovation, we created *Observer*, a multimodal dataset comprising videotaped outpatient visits, electronic health record (EHR) data and structured surveys. This paper describes the data collection procedures and summarizes the clinical and contextual features of the dataset.

**Materials and Methods:** A multistakeholder steering group shaped recruitment strategies, survey design, and privacy-preserving design. Consented patients and primary care providers (PCPs) were recorded using room-view and egocentric cameras. EHR data, metadata and audit logs were also captured. A custom de-identification pipeline, combining transcript redaction, voice masking, and facial blurring, ensured video and EHR HIPAA compliance.

**Results:** We report on the first 100 visits in this continually growing dataset. Thirteen PCPs from four clinics participated. Recording the first 100 visits required approaching 210 patients, from which 129 consented (61%), with 29 patients missing their scheduled encounter after consenting. Visit lengths ranged from 5 to 100 minutes, covering preventive care to chronic disease management. Survey responses revealed high satisfaction: 4.24/5 (patients) and 3.94/5 (PCPs). Visit experience was unaffected by the presence of video recording technology.

**Discussion:** We demonstrate the feasibility of capturing rich, real-world primary care interactions using scalable, privacy-sensitive methods. Room layout and camera placement were key influences on recorded communication and are now added to the dataset. The *Observer* dataset enables future clinical AI research/development, communication studies, and informatics education among public and private user groups.

**Conclusion:** Observer is a new, shareable, real-world clinic encounter research and teaching resource with a representative sample of adult primary care data.

## Background

The U.S. healthcare system—once a global exemplar—is now criticized for its high costs and relatively poor health outcomes.^1^ While biomedical researchers and industry leaders have proposed incremental improvements to existing technological infrastructure—once seen as key to reform^2–4^—these efforts have proven inadequate for addressing the deeper systemic issues. Johnson refers to this as the “Era of Entanglement,” a period marked by the intertwined demands of clinical care, regulation, and finance that have produced a fragmented, burdensome system.^5^ As with the leap from horse-drawn carriages to motorized vehicles, transformative innovation, rather than iterative, is needed to advance healthcare delivery meaningfully.

The National Research Council defines this thinking as radical change, where healthcare exploits “qualitatively new ways of conceptualizing and solving health and healthcare problems and revolutionary ways of addressing these problems.”^4^ However, these researchers must understand how healthcare works — they need access to data and observations that spur new hypotheses and ideas. The conundrums they face are the challenges our current environment presents for non-medical faculty to obtain access to patient care settings, due to hospital and privacy policies.

Video as a data collection tool has been underutilized in healthcare due to inherent barriers, including confidentiality, privacy, and the technical challenges associated with sharing large file sizes.^6–8^ Earlier efforts to capture clinical visit knowledge recognized that tacit knowledge, non-verbal interpersonal interactions, and the necessity of understanding ‘embodied cognition’ all emphasize the need to video-capture interactions between physicians and their patients in a natural setting.^6,9–11^ These data also enable the documentation of activities in their entirety and complexity.^12^ It builds on the work of others who have used videotapes and eye-tracking methods. ^13–15^: Video capture alone, however, is an incomplete source of clinic visit details, due to the increasing role of electronic health records, web applications, and other in-room technologies.^16^ A systematic review of 175 studies found that only 11% examined the role of computer or electronic medical record use during the encounter.^7^ However, there is at least a 35-year corpus of studies of primary care visits that combine qualitative and quantitative research methods.^16–25^ While these did not result in systematic databases, they often stressed the need for such structures.^26–28^ The National Academies Press published a major compendium of these efforts.^16^

Combining video data with a view of the activities on the computer (EHR logs, clinical notes, and visit-related surveys) as well as room layouts showing the position of the technology with respect to the patient provides a multifaceted view, allowing for qualitative and quantitative analysis and a view supporting a holistic analysis of encounters. This perspective is especially intriguing given current advances in multimodal AI.^29–35^

There are also concerns that the process of video recording may bias the encounter, thereby lessening the value of these recordings due to the Hawthorne effect.^36^ Interestingly, video recordings are shown to be less disruptive to natural participant behavior than human observers present during visits.^6^ Another study examining physicians’ consultation behavior found no evidence that awareness of video recording affects objective measures of their performance, supporting the use of videos as valuable tools for teaching and research.^36^

To address the challenges of researcher access to clinic encounters and to embrace the need for multimodal data, we created the Observer dataset, a multimodal resource for multidisciplinary research. This paper describes the efforts and observational frameworks, as well as initial studies and observations, enabled by the dataset.

## Materials and Methods

The overarching goal of Observer is to provide a library of clinical exam rooms and interactions among patients, any other caregivers, the primary care provider (PCP), and technology. Our goal is to provide a downloadable dataset for users interested in research directly related to clinical exam rooms. We anticipated that researchers, educators, and policymakers would be the primary audience for this resource. Ultimately, our plan is for Observer to be housed in a repository that adheres to FAIR principles^37^ as defined below:

- *Findable* by providing metadata that clearly describes the data using a dashboard, being indexed in searchable repositories, and using a digital object identifier to provide a persistent link to its internet location.
- *Accessible* by providing the website URL (https://observer.med.upenn.edu/) and data availability statement on a series of public sites, in addition to our lab site.
- *Interoperable* by mapping medical data to standard vocabularies using the OMOP table schema for Observer data. *Reusable* by providing clear usage licenses and documenting information provenance, including any sources for derivative metadata.

The remainder of this paper describes the dataset creation process and initial observations. Of note, this paper focuses on our first 100 recorded visits. We continue to record visits, intending to collect 200 visits in adult primary care settings by the end of 2026.

### Design Considerations

We began designing Observer by consulting with two steering groups of adult primary care stakeholders. The first consisted of adult patients and family members. Following a brief presentation by the PI, the group responded to a series of driving questions. They were all very supportive of the project and excited about how it might be used to improve their interactions with physicians and advanced practice nurses. They made the following recommendations for our recruitment and data collection plans:

1. Identify that we are Penn Medicine researchers working with their clinic team (and not, for example, billing or marketing personnel).
2. Compensation for time and effort should be in the form of a $50 physical gift card, rather than one that requires a bank account or an email address.
3. Allow the patient to wear an egocentric (forehead-mounted) camera. Our initial plan was only to ask the PCP to wear this camera, to provide eye tracking as well as a view of the EHR screen. This focus group suggested that observing the patient’s experiences would be invaluable.

Strongly consider asking patients and their families for feedback on their visit. Our initial plan was only to ask clinicians for feedback, but patients immediately recognized our plan did not include them, which might cause us to miss their valuable insights. We agreed to incorporate a post-visit patient survey. A second set of focus groups consisted of clinic physicians and nursing staff. After a short presentation by the PI, the group discussion led to the following recommendations.

1. Consider using an after-visit clinician survey instead of requiring clinicians to review their visit recordings and comment on the visit. Clinicians were uncomfortable seeing themselves on the video.
2. To minimize clinic disruption, begin by recording only a few visits for each consented clinician, and then progress to recording multiple visits after staff and consented clinicians are comfortable with the processes.
3. To avoid the risk of new patients losing trust in the clinic team, confirm which patients we would approach until the consented clinicians and clinic managers were comfortable with the study processes.

Based on these conversations and a review of similar medicine-related data platforms,^13–15,38–40^ we constructed a data plan for Observer that holistically covered the activities of a typical ambulatory care visit. We subsequently obtained IRB approval for visit recording and data collection using the process described below.

As noted, this dataset consists of ambulatory care visit videos linked to the EHR user action log, encounter summary, current and past diagnoses, problems, medications, allergies, and all visit-related correspondence from 24 hours before until 48 hours after the recorded visit. Transcripts and final progress notes accompany each of these videos.

Our review of existing efforts to create ambulatory visit data sets was disappointing. In particular, none of these platforms supported the retention of multimodal PHI, which requires sophisticated tools for removing identifiers across modalities (facial recognition blurring, voice alteration, and metadata scrubbing) that go far beyond standard text-based de-identification ^41,42^ and extensive curation. Therefore, we realized that to share these data, we would need to create a de-identification pipeline, described below.

### Data Model

To support interoperability with existing EHR-based data and less commonly available video-derived data, we developed a custom data model inspired by OMOP and MIMIC to structure our EHR-derived fields, visit metadata, and survey instruments.

### Equipment Selection

We tested numerous cameras, microphone setups, and screen capture software tools under simulated conditions before selecting our current equipment for collecting encounter information. After tests and discussions with health system security experts, we ruled out using screen-capture software due to the difficulty in aligning those data with room videos, challenges with some software capturing video from more than one application (such as from both the EHR and a web browser) and other equipment challenges, (e.g., bringing in laptops with enterprise licenses for PCPs to use during the visit).

After testing various configurations, we selected the following:

- Room-view camera with a microphone array and a 360-degree room view, connected to a local Zoom session, monitored by a research assistant outside the room.
- Camera worn on the head at eye level, providing both a microphone and a video from the wearer’s perspective (egocentric view), offered to both the PCP and the patient.
- For telemedicine encounters, we selected Zoom, which supported secure video downloads, and with which clinical teams and patients were already familiar.
- To standardize and facilitate remote consent for patients, we initially used an established secure messaging platform.

We also learned that linking EHR use to the line-by-line transcripts of PCP-patient interactions is a valuable alignment for end users interested in understanding, for example, what is being discussed while the computer is in use.

Previous approaches to video-capturing clinical interactions, such as those by Montague and Asan,^43^ relied on multiple fixed cameras positioned at different angles to capture gaze direction. We instead incorporated egocentric head-mounted cameras for both patients and physicians to capture first-person perspectives of what each participant sees during the visit. We also built on the previous work of others in this space, including Senathirajah and colleagues,^13^ whose scoping review confirmed that automated detection of eye gaze without eye tracking was feasible and adequate; and Khairat and colleagues,^14^ who found that user experience can be measured using surveys for physician satisfaction, system usability, and workload intensity. These latter observations motivated the design of our physician survey. More technical details may be found at https://observer.med.upenn.edu/.

### Recruitment and Data Collection Process

Our study recruitment focused on patients and PCPs in adult primary care clinics throughout the University of Pennsylvania Health System, an integrated delivery system. We adopted the suggestion of our multistakeholder steering group and required that PCPs be enrolled first, followed by a convenience sample of their patients. Our final recruitment strategy is below and summarized in Supplemental Figure 1. All recruitment materials and any changes made in our protocol are available at https://observer.med.upenn.edu/.

#### Primary Care Provider Enrollment

We presented the project’s goals to primary care providers during their weekly team meetings. After the presentation and question-answering, we provided informed consent forms (ICFs) to each PCP. The ICFs also included options for the degree of de-identification each PCP required for broad consent. Initially, consented PCPs were contacted to establish their preference for days and times best suited for video recording. This process was relaxed once PCPs and we became more comfortable with the study protocol.

#### Patient Enrollment

Our study team contacted any patients who were scheduled for visits within consented PCP’s date and time preferences, described the study, and reviewed the consent form using an IRB-approved script. Patients who were willing to participate then received a link to the consent form via email or text message with an option to electronically sign the form or were met by our team to review and sign the ICF before the visit. As offered to providers, patients also had the option to blur their face and/or obscure their voice, with an additional option to blur any other specified body parts. Patients were not recorded if ICFs were not signed before any actual visits. The University of Pennsylvania IRB approved this process.

#### Exam Room Preparation and Visit Data Collection

Before visits, a research coordinator set up in-room cameras, then met patients in the room to help them place and activate the egocentric camera. Any additional people accompanying the patient signed an informed consent to be recorded. They had the option of blurring their image and audio. After visits, the research coordinator collected all recording equipment and asked patients to complete the short survey about the visit while still in the room (under 5 minutes on average). Once the surveys were completed, the research coordinator provided patients with gift cards and escorted them out of the exam room. Primary care providers were given an exit survey to complete and return to the research team. They received a gift card after returning the exit survey.

#### EHR Data Extraction

After the visit, a research coordinator completed collecting all data for the visit. Recorded videos were copied to a secure cloud platform (Microsoft Azure Databricks) for de-identification and removal of extraneous events, such as turning on the video recorders, which were already captured by the recorders. Survey results were labeled with a visit ID and stored electronically in a secure location. A request was made to our data and analytics core to extract all pertinent data from the EHR and to label these data with the same visit ID. These data were provided to us without PHI and with dates shifted. We updated all data to reflect this new date and ID and stored these data in our same secure location. All other data derived from the visit video were managed by us, with labels provided by our honest broker. All other labels were removed before the data were available for review.

### Privacy Preservation

To preserve privacy, we developed and implemented MedVidDeID,^44^ a modular pipeline designed to de-identify clinical audio-video (AV) data in compliance with HIPAA Safe Harbor standards.^45^ This system integrates state-of-the-art, open-source tools into a six-stage workflow that treats audio and video modalities independently. First, we extract transcripts using WhisperX and de-identify them with a local instance of PHIlter. Audio was de-identified by redacting timestamped PHI segments and transforming voice characteristics to obscure speaker identity. Video frames are processed using YOLOv11 to detect and blur facial features, utilizing pose estimation and Kalman filtering. The de-identified audio and video are then recombined and subjected to manual quality control to ensure completeness of PHI removal. An analysis of our first 11 hours of clinical encounters demonstrated a 91% video de-identification rate, reducing the need for manual processing time by over 60%. This package enables scalable, secure sharing and analysis of AV clinical data while preserving patient privacy.

## Results

### Participant Recruitment

We recruited 13 PCPs from 4 clinic settings. We approached 210 patients scheduled to see these PCPs to participate in this study. A total of 129 patients agreed to participate (61%), from which we were able to capture clinic visits for 100 (78%). There were 29 patients who agreed to participate but were not recorded due to illness, rescheduling, or logistical issues. Three patients participated more than once for separate visits. We contacted three patients whose primary language was not English, of whom two participated in the study (data not shown). Figure 1 summarizes enrolled patients self-reported health and educational levels.

**Figure 1.**
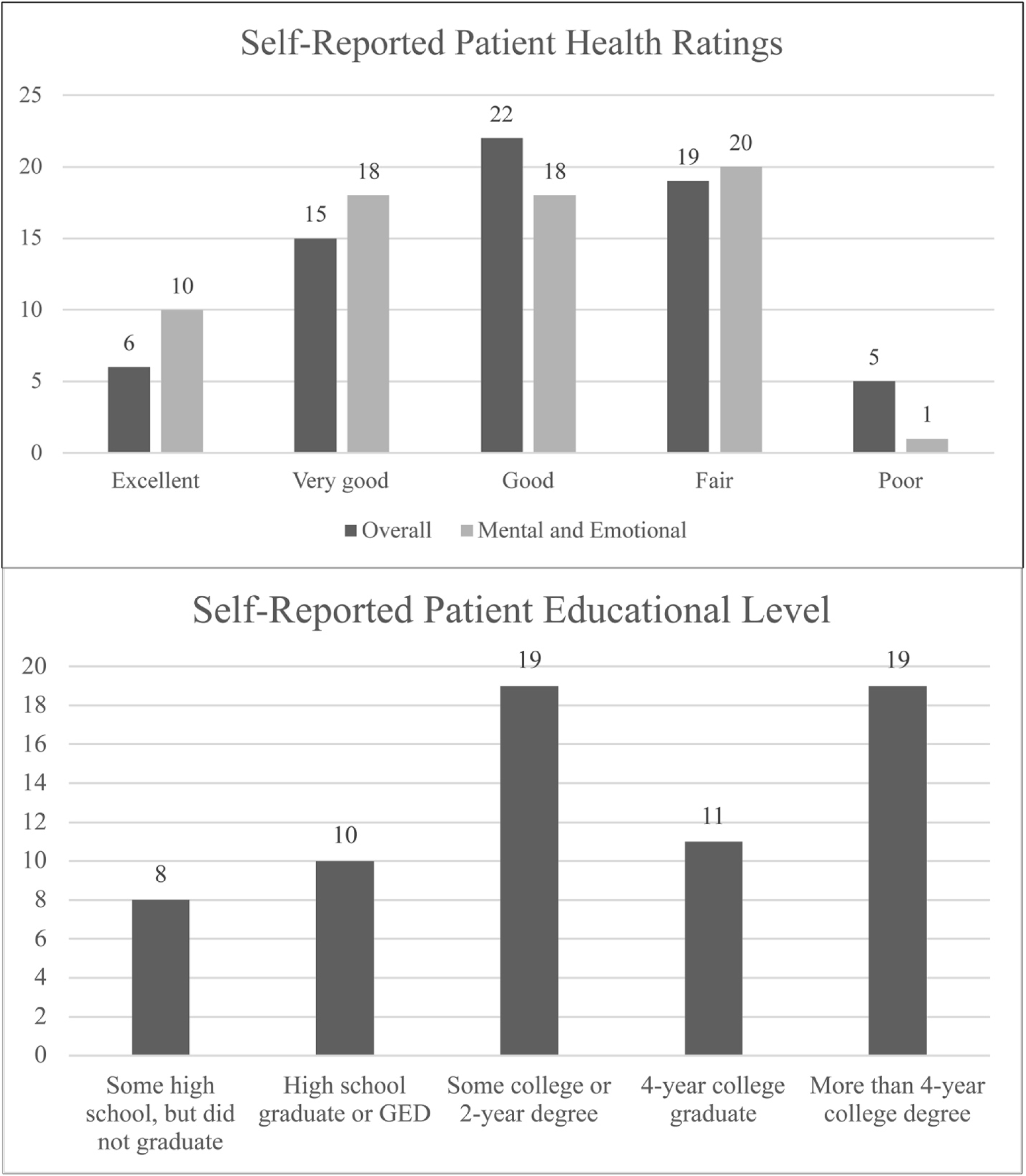
Patient characteristics. Data only available for the last 70 visits, due to the addition of these questions on the post-visit surveys.

Tables 1 and 2 summarize the results of our first 100 visits. Visit types, as extracted from EHR data, represent the range of visit types for primary care.

**Table 1:**
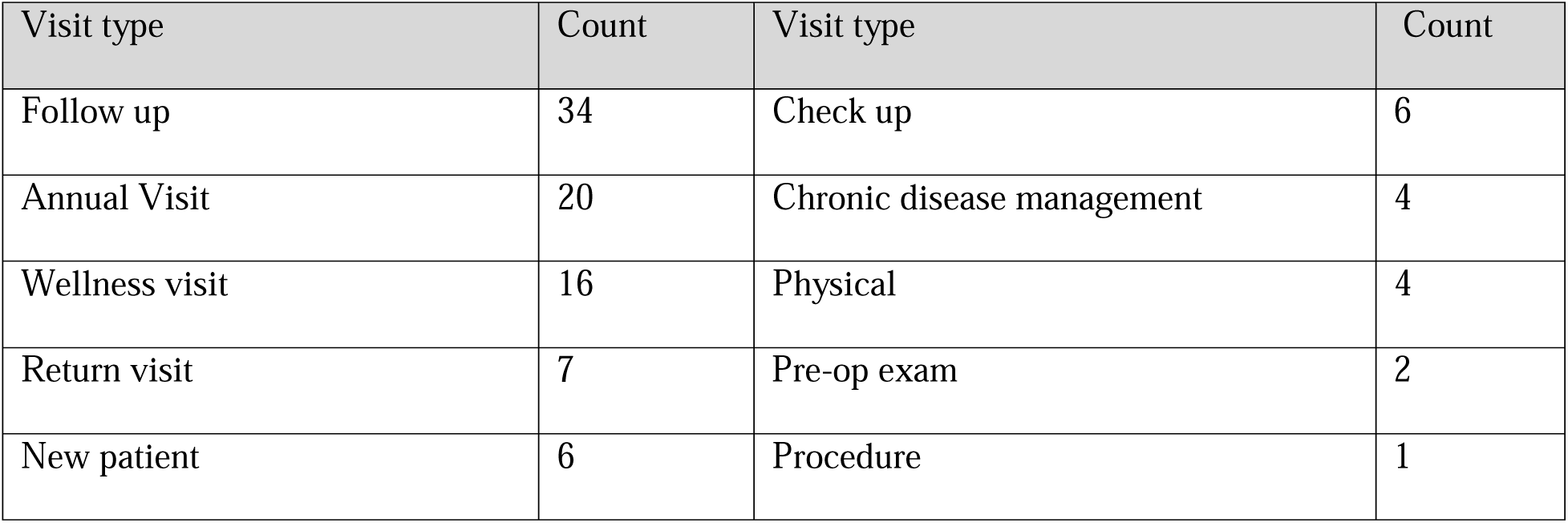
Visit characteristics.

| Visit type | Count | Visit type | Count |
| --- | --- | --- | --- |
| Follow up | 34 | Check up | 6 |
| Annual Visit | 20 | Chronic disease management | 4 |
| Wellness visit | 16 | Physical | 4 |
| Return visit | 7 | Pre-op exam | 2 |
| New patient | 6 | Procedure | 1 |

**Table 2.**
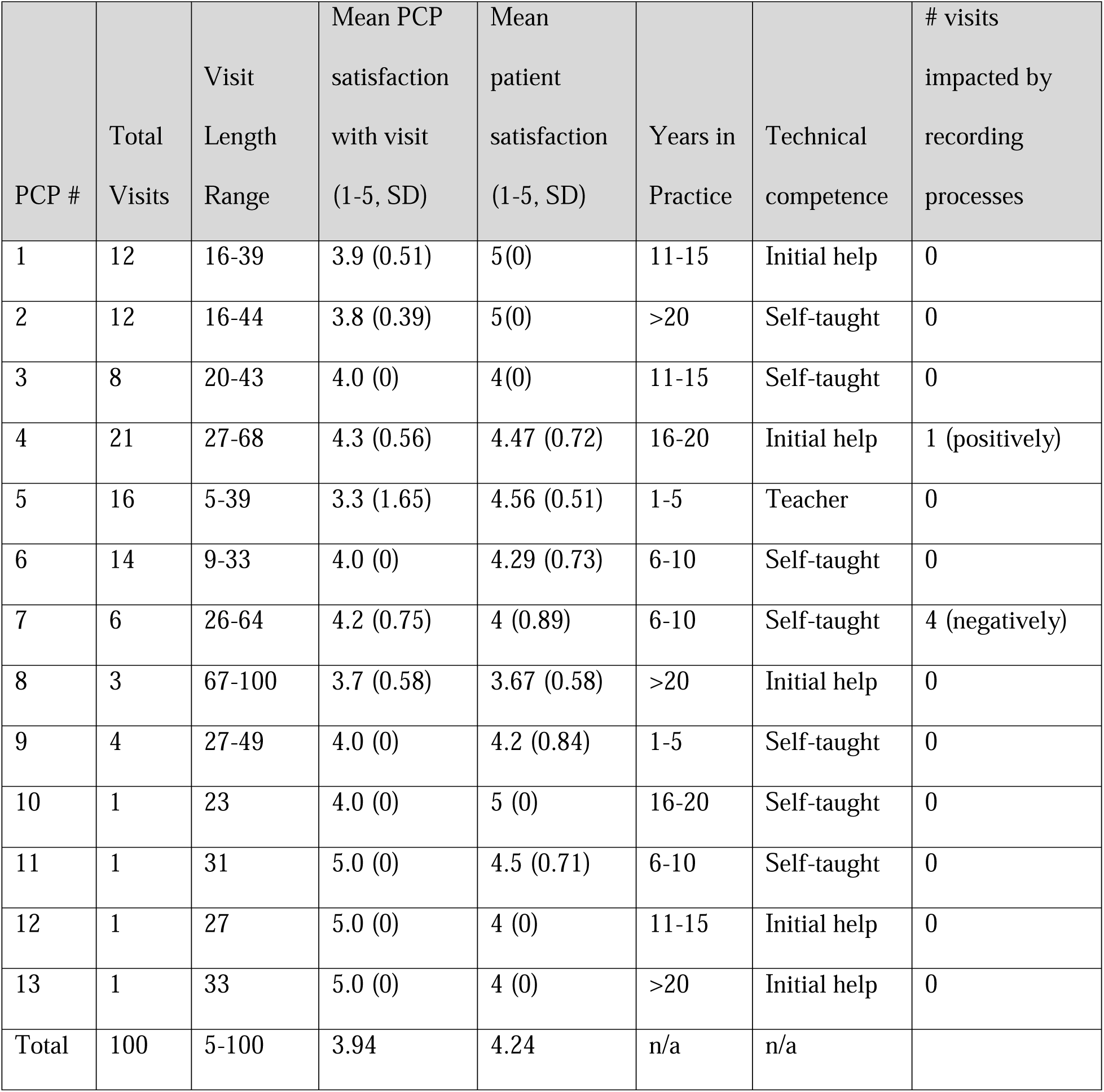
Data about primary care provider (PCP) characteristics were obtained via self-reported post-visit surveys Possible answers to the self-reported technical competence question (“How would you rate your comfort level with using technology in healthcare?”) ranged from being a novice (“Initial help”) to being comfortable with technology (“Self-taught”) to being considered an expert (“Teacher”).

| PCP # | Total Visits | Visit Length Range | Mean PCP satisfaction with visit (1-5, SD) | Mean patient satisfaction (1-5, SD) | Years in Practice | Technical competence | # visits impacted by recording processes |
| --- | --- | --- | --- | --- | --- | --- | --- |
| 1 | 12 | 16-39 | 3.9 (0.51) | 5(0) | 11-15 | Initial help | 0 |
| 2 | 12 | 16-44 | 3.8 (0.39) | 5(0) | >20 | Self-taught | 0 |
| 3 | 8 | 20-43 | 4.0 (0) | 4(0) | 11-15 | Self-taught | 0 |
| 4 | 21 | 27-68 | 4.3 (0.56) | 4.47 (0.72) | 16-20 | Initial help | 1 (positively) |
| 5 | 16 | 5-39 | 3.3 (1.65) | 4.56 (0.51) | 1-5 | Teacher | 0 |
| 6 | 14 | 9-33 | 4.0 (0) | 4.29 (0.73) | 6-10 | Self-taught | 0 |
| 7 | 6 | 26-64 | 4.2 (0.75) | 4 (0.89) | 6-10 | Self-taught | 4 (negatively) |
| 8 | 3 | 67-100 | 3.7 (0.58) | 3.67 (0.58) | >20 | Initial help | 0 |
| 9 | 4 | 27-49 | 4.0 (0) | 4.2 (0.84) | 1-5 | Self-taught | 0 |
| 10 | 1 | 23 | 4.0 (0) | 5 (0) | 16-20 | Self-taught | 0 |
| 11 | 1 | 31 | 5.0 (0) | 4.5 (0.71) | 6-10 | Self-taught | 0 |
| 12 | 1 | 27 | 5.0 (0) | 4 (0) | 11-15 | Initial help | 0 |
| 13 | 1 | 33 | 5.0 (0) | 4 (0) | >20 | Initial help | 0 |
| Total | 100 | 5-100 | 3.94 | 4.24 | n/a | n/a |  |

PCPs were uniformly distributed across years in practice, ranging from 1 to 5 years to more than 20 years in practice. In response to the question, “How would you rate your comfort level with using technology in healthcare?” most providers described themselves as either self-taught or needing instruction (see Table caption for definitions.) Visit length ranged from 5 to 100 minutes. PCPs rated their visit satisfaction as a weighted average of 3.95/5 across all visits, while their patients rated visits as 4.24/5 on average, with occasionally discrepant scores provided by the patient and clinician for the same visit (data not shown, but available on the Observer website). Overall, the visit experience was unaffected by the presence of video recording technology. Of those who perceived some impact, one was positive, where the PCP performing a physical exam reminded a medical student that it was always important to examine the patient without a shirt on. Another PCP reported a negative impact without specifying details.

### Observer Composition and Structure

At the time of this publication, the Observer dataset comprises 110 outpatient primary care visits, each recorded using a standardized protocol. For each visit, Observer includes:

- **Room-view video recordings**, captured with a stationary 360-degree camera equipped with a directional microphone array, are positioned to provide a wide-angle perspective of the physical interaction between patient and clinician.
- **Egocentric videos**, recorded using head-mounted cameras worn by patients and PCPs. These recordings offer a first-person perspective of visual attention, body language, and interpersonal engagement. They also provide the equivalent of eye-tracking, enabling a view of how computer technology is used during visits.
- **EHR-derived audit logs**, including timestamped records of all user interactions within the electronic health record during the visit and the surrounding 72-hour window (24 hours before through 48 hours after the encounter).
- **Structured metadata**, covering reason for visit, patient demographics, visit type, clinician characteristics, room configuration, and survey responses.
- **Survey results** from patients and clinicians after the visit, capturing perceptions of communication quality, comfort, and the perceived impact of recording technology.
- **Transcripts**, automatically generated and manually verified, reflecting verbal exchanges between participants.
- **Clinical notes,** including timestamped text entries from providers linked to specific patient visits, capturing detailed observations, diagnoses, and care plans, with metadata on note type and status.
- **Vital signs and measurements,** recorded during visits, including quantitative data such as blood pressure, weight, height, pulse, and oxygen saturation, linked to the corresponding patient encounter.

To support scalable reuse, we provide standardized file formats (MP4 for video, CSV for logs and metadata, and JSON for annotations) along with accompanying documentation that describes data provenance, schema definitions, and known quality attributes. All data packages are stored in a secure institutional archive, with download access granted through a verification process modeled after the MIMIC framework.

### Clinic Encounter Visit Details

Figure 2 summarizes the visit diagnoses covered in our first 100 recordings. Patients had multiple active diagnoses with a frequency representative of a typical adult primary care setting.

**Figure 2.**
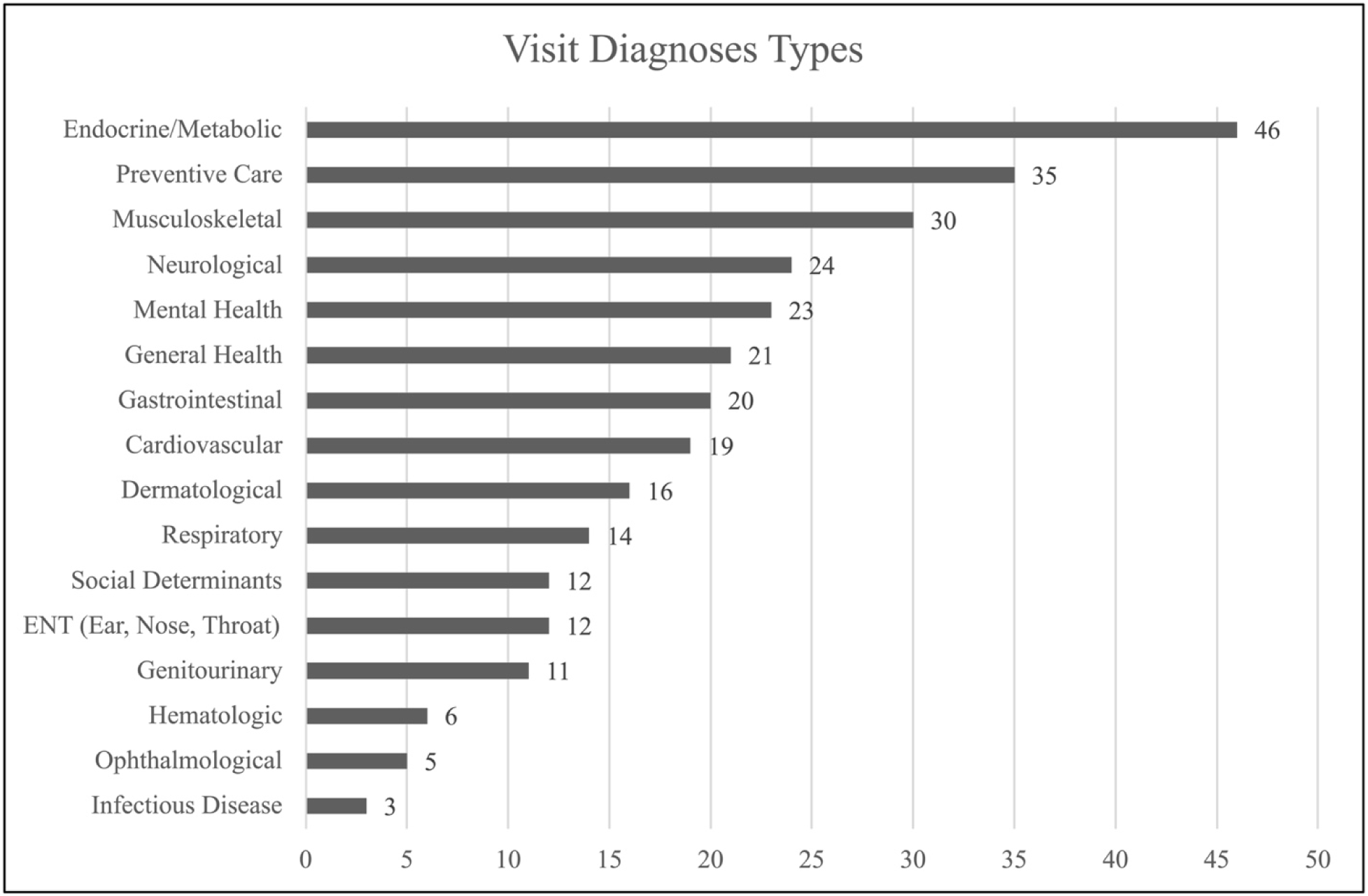
Summary of visit diagnoses.

Figure 3 provides a sample of the different room configurations. We observed that the physical configuration of exam rooms and the EHR screens significantly affects the ability of PCPs and patients to see each other face-to-face and to view the EHR.

**Figure 3.**
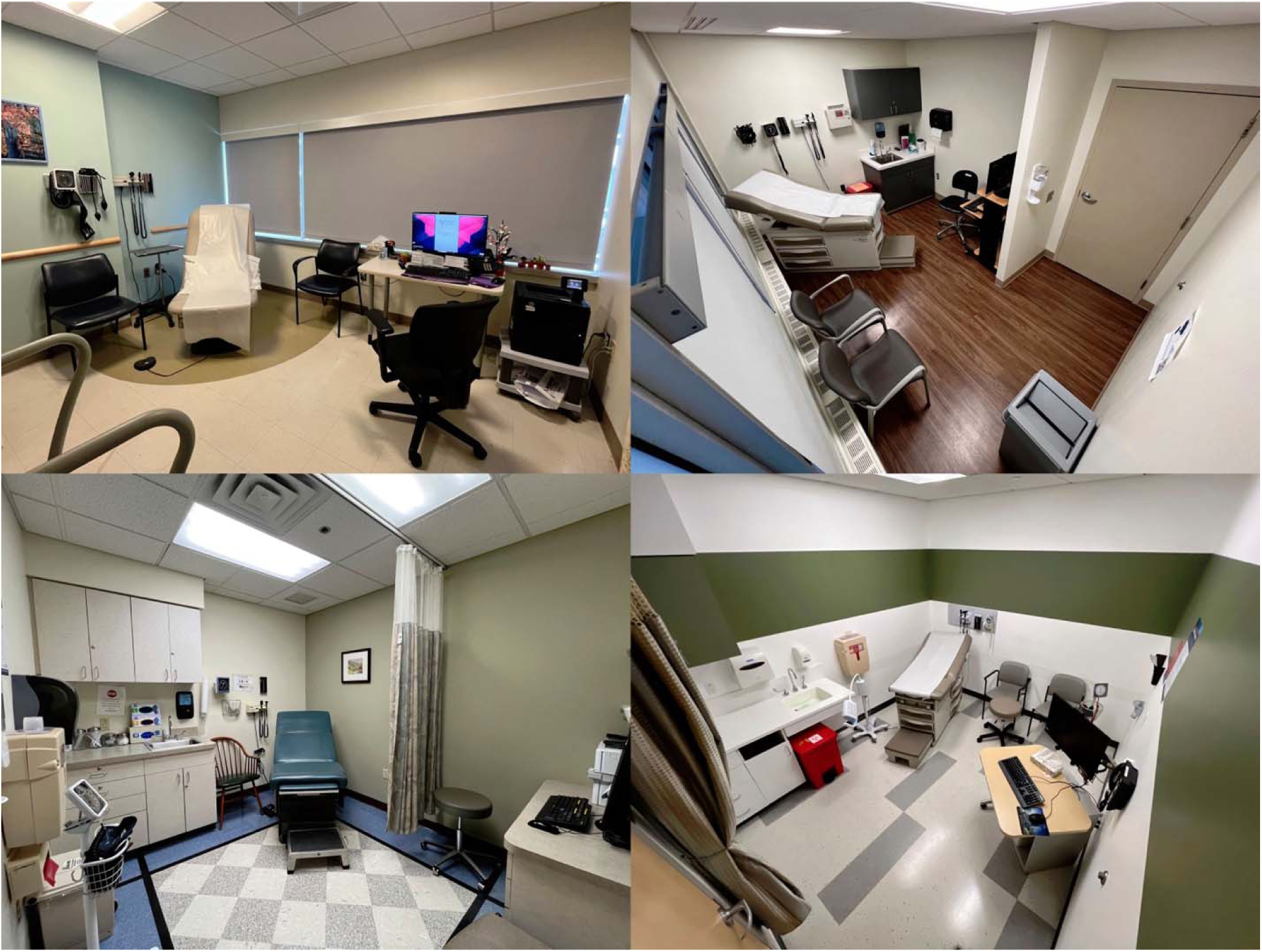
Sample room configurations of participating clinical sites.

## Discussion

We report our experience in creating the Observer dataset — a first-of-its-kind library of visit recordings, EHR data, and various other forms of accompanying information. Previous efforts, such as the Activity Log Files Aggregation toolkit, while methodologically sound, lacked the infrastructure for long-term data sharing and became inaccessible over time, requiring subsequent studies to collect data independently rather than build upon existing foundations —a limitation that Observer aims to address. We anticipate Observer will enable hypothesis generation and testing, as well as an evolving, valuable dataset for model development and testing. Ethnographic researchers will be able to use Observer to evaluate compliance with healthcare delivery recommendations, observe care provider-patient interaction, and identify opportunities to improve both needed resources and cognitive support for both patients and PCPs.

Room configuration varied significantly across different clinics and is likely an essential covariate in studies of communication, as noted by Kumarapel^46^ and summarized in a systematic review.^47^ For this reason, we added an image of the room configuration and metadata about this configuration to each visit’s data set. These videos provide natural experiments to observe how screen and room arrangements affect the PCPs’ time spent looking at EHR screens versus looking at patients, and how PCPs utilize EHR data in dialogues and explanations.

In addition to hypothesis generation and testing,^3^ these recordings allow a complete record of consultations to be viewed repeatedly for inter-observer reliability testing and open science.^48^ Following discussions with researchers at recent meetings, we believe the multimodal nature of Observer can be beneficial for the classes of problems outlined, as examples in Table 3 (below).

**Table 3.** Example Use Cases for the Observer Dataset and Their Required Data Modalities.

| Use Case | Example | Video | Transcript | EHR data | Audit Log | Room Audio |
| --- | --- | --- | --- | --- | --- | --- |
| Ethnographic Research | A researcher could compare visits with and without an automated scribe to measure differences in the frequency of direct patient-provider eye contact and the number of conversational interruptions. | X | X |  |  | X |
| Medical Education | A medical educator could task students with watching an Observer video and using a standardized checklist to evaluate the PCP's communication skills, then discussing the findings as a group. | X | X |  |  | X |
| Visit Summarization | A software engineer could train a model to generate a clinical summary from the visit transcript and then validate its accuracy by comparing the output against the physician's official EHR note. |  | X | X |  | X |
| Quantify Signs and Symptoms | A data scientist could develop a model that analyzes features (e.g., pitch, speaking rate, pauses, gait) to create a quantitative score for cognitive impairment, correlating it with relevant EHR data. | X | X | X |  | X |
| Cognitive Support Assessment | A human factors researcher could identify points where a PCP accesses a clinical guideline and analyzes subsequent dialogue to find how that information was raised in the patient conversation. | x | x |  | x | x |
| Administrative Burden Assessment | A health services researcher could quantify the amount of in-visit time wasted on administrative tasks (e.g., verbal discussion about next visit, time spent scheduling next visit) versus clinical assessment. |  | x | x | x | x |
| Visit summarization | Fine-tuning a language model to generate patient-friendly visit summaries. |  | x | x |  |  |
| Cognitive health monitoring | Detecting speech markers of mild cognitive impairment across longitudinal visits. |  | x | x |  | x |
| Predictive modeling (ML) | Predicting patient no-shows based on pre-visit behaviors and clinician communication patterns. | x | x | x | x | x |
| New technology evaluation | Assessing usability of an AI-powered voice assistant for automated documentation in exam rooms. | x | x | x | x | x |

A key contribution of the Observer dataset is the ability to synchronize observational data with process data extracted from EHR logs. These data can serve as powerful new sources for Audit and Feedback (A&F), as they help generate the actionable plans with specific advice for improvement that are known to make feedback most effective.^49^ This dataset, along with its ability to generate specific and actionable feedback, helps catalyze multimodal AI applications in various contexts. For example, we are developing a model for identifying agenda topics using de-identified visit transcripts from Observer.^50^ Another team is using multimodal data to develop feature-detection models that predict mild cognitive impairment. These data also support model fine-tuning and provide examples for few-shot prompting of large language models. Other contexts will likely involve clinical decision support rules, providing real-time visit information, an almost infinite array of clinical care improvements, physician training, the use and design of EHRs, the role of AI, patient-facing chatbots, and exam room redesign.

### Limitations and Challenges

Although we envision many uses for Observer, we recognize that any data resource of this type suffers from limitations that are either measurable or implied. For example, we have observed no systematic skew in recruiting physicians or patients; however, physician liability concerns could introduce a selection bias in their participation, resulting in a biased sample of patients invited to participate. These potential biases necessitate vigilant monitoring and the implementation of measures to ensure the collection of representative samples. The ethical implications of video recording and the possible risks to participant confidentiality and privacy must be meticulously addressed to maintain the integrity of the research and the trust of the participants. Balancing the benefits of rich, detailed data collection with the ethical considerations and participants’ comfort is crucial in leveraging representative video recordings for research purposes.^51,52^

The request for video recordings may have raised concerns among potential participants. Although we did not collect data from non-respondents, some individuals may have been reluctant to participate because of the possibility that a) multiple individuals may view the video recordings over time, b) unauthorized individuals may access video data due to improper storage, and c) an individual’s identity could be more easily discerned from a video recording than from written data. Although we emphasized privacy protections and included clear language about participants’ right to withdraw at any time, some patients still expressed concerns that influenced their willingness to participate. We provided modest gift cards as a token of appreciation, and we allowed patients to select the level of privacy preservation they required to participate, as part of the informed consent process. These strategies seemed to support participation and may be helpful for others collecting encounter video data.

## Conclusion

Observer builds on previous work to create a data resource containing a completely de-identified view of ambulatory care visits—with multiple camera views, a visit transcript, an EHR audit log covering from 24 hours before the visit until 48 hours after the visit, and all pertinent EHR data used or accessible during the visit. Creating the Observer dataset required attention to a holistic view of a clinical encounter, as well as ensuring patient and PCP privacy using state-of-the-art techniques. Next steps will focus on developing the dataset into a repository available for general use.

## Data Availability

All data produced are available online at https://observer.med.upenn.edu/

https://github.com/kbjohnson-penn/observer_frontend

## Data Availability

The Observer dataset is available at https://observer.med.upenn.edu/. Requests for access to its data may be made by following the instructions on that website.

## Code Availability

All code related to this project is available on GitHub, at https://github.com/kbjohnson-penn/observer_frontend

## Acknowledgements

We appreciate the thoughtful review of this manuscript by Dr. S. Trent Rosenbloom and Dr. John Holmes, as well as the proofreading of this manuscript by Lauren Malloy.

## Author Contributions

All authors contributed to the conception and design of this project. Mr. Mopidevi provided the technical work leading to this manuscript and contributed to the writing of the Methods section. Mr. Alasaly and Dr. Johnson analyzed the data reported in this manuscript. Drs. Koppel, Jang, and Eaton contributed to the interpretation of the data reported in this manuscript. All authors approve the submitted version of this manuscript.

