## Supplementary figures and images for "Observer: Creation of a Novel Multimodal Dataset for Outpatient Care Research"

### Supplemental Figure 1

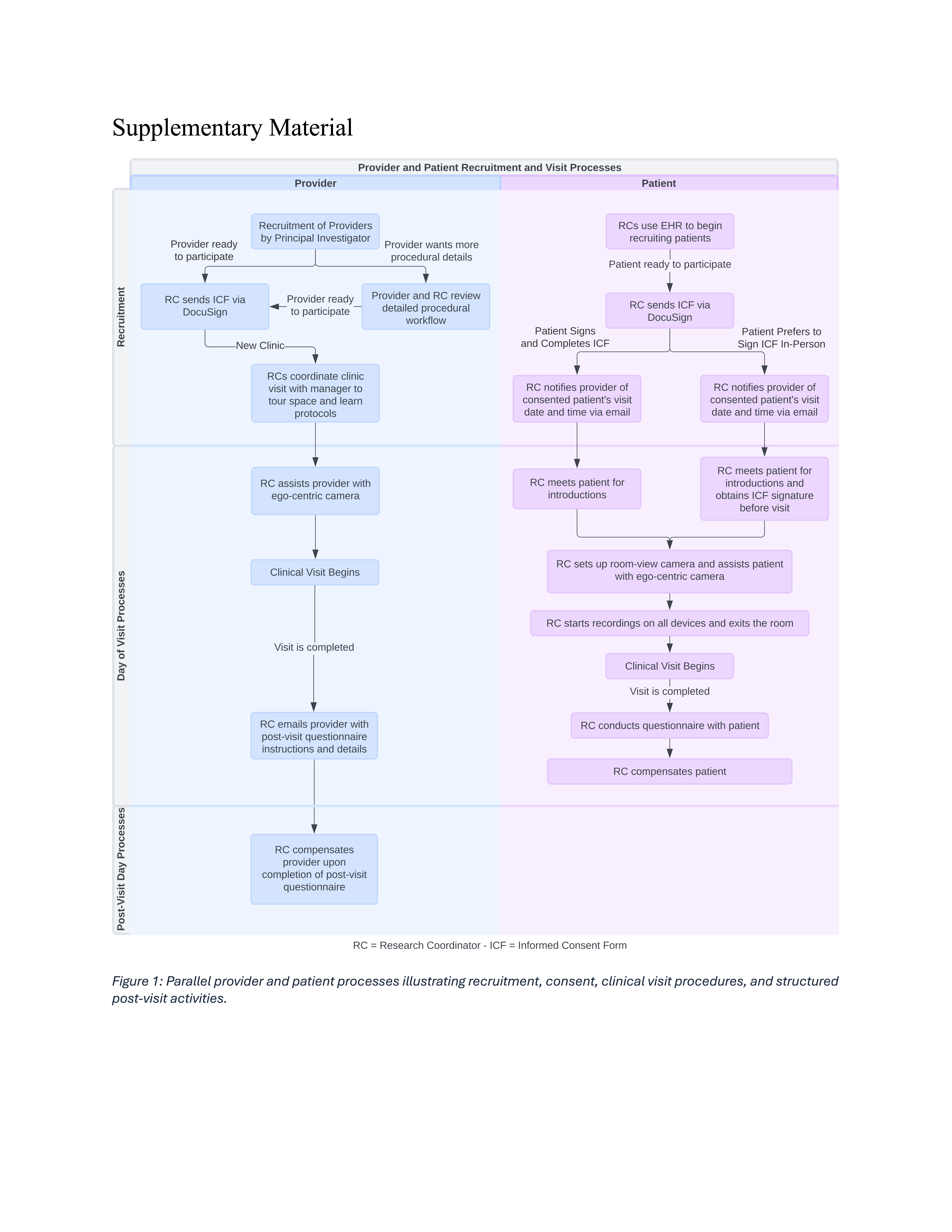
